# Application and reflection of blended learning in theoretical and practice courses of periodontology education

**DOI:** 10.1101/2022.04.22.22274171

**Authors:** Jiachen Dong, Yue Liao, Xia Cao, Huiwen Chen, Zhongchen Song

**Affiliations:** Department of Periodontology, Ninth People’s Hospital, School of Medicine, Shanghai Jiao Tong University, Shanghai, China; Shanghai Key Laboratory of Stomatology, Shanghai Research Institute of Stomatology, Shanghai, China; National Clinical Research Center of Stomatology, Shanghai, China; College of Stomatology, Shanghai Jiao Tong University, Shanghai, China

**Keywords:** Blended learning, Online learning, Periodontal education, Undergraduate medical education

## Abstract

**Background:** Blended learning mode has been widely applied in medical education, especially during the current COVID-19 pandemic. This study is to evaluate the effectiveness of blended learning mode in periodontal education for further promotion.

**Methods:** The blended learning mode consists of face-to-face classes and SPOCs built on the Xfaike online course platform. The preparation before the courses included the optimization of teaching calendar, establishment of online learning platform, integration of the online and offline content. The feedback was evaluated by a questionnaire involving instruction method, instruction content and learning outcomes.

**Results:** All of the participants (n=65) answered the questionnaire. According to the questionnaire, the blended learning mode has been approved by all of the students. More than 95% of students convinced that blended learning mode can improve their self-learning ability. About 84.62% of students could finish the preview and review task of online courses. All of the students believed that they can exchange their questions with teachers more pertinently in offline classes after online learning, and online learning videos can be viewed smoothly and repeatedly at any time.

**Conclusions:** The blended learning mode has been highly accepted among students, as the learning platform could provide abundant learning materials, improve self-learning ability, and provide a deeper understanding of knowledge. Thus, the blended learning could be considered as a promising educational mode for medical students to meet higher educational requirements.

## INTRODUCTION

Periodontology, as a required course for dental students, proposes high demand on both theoretical knowledge and practical skills. The teaching method of dental education has been changed constantly to meet the requirements for the cultivation of dental talents.^1^ Therefore, the teaching methods have also been optimized in our school with the application of blended learning.

Blended learning consists of both online learning and face-to-face offline learning, which has been widely applied in medical education.^2,3^ Compared with traditional offline learning format, blended learning demonstrated better effects on knowledge outcomes consistently, which is a promising option for medical teaching.^3,4^ The blending learning retains effectiveness of traditional offline class. Meanwhile, online learning allow the students to have collaboration and discussion without the limits of time and space, and develop individualized learning program.

Online learning, also called e-learning, is booming with the development of internet.^5^ Numerous e-learning programs are involved in medical and dental education.^6^ When compared with offline learning, online learning could widen the horizon and knowledge of audiences, and give access to latest scientific achievements.^7^ The reformation of online education was witnessed in 2012 owning to the massive open online courses (MOOCs).^8^ Even though millions of students have benefited from MOOC, and MOOC made up for shortages of traditional university teaching, it still faces many challenges. Considering the massive audiences of MOOC, the imbalance of teacher-student ratio is inevitable, leading to low student attendance and completion rate of courses.^9^ What’s more, students are unable to get enough attention and opportunities to communicate with teachers.

In 2013, Professor Armando Fox from University of California proposed a small private online courses (SPOCs), which could also provide abundant educational resources and avoid many the management problems, Thus, small private online course (SPOC) has been popular in medical education with some positive feedback, as it can meet the requirement for further interaction and communication in professional courses.^10,11^ Besides, the combination of SPOC and offline class has already been carried out in some medical school. The blended teaching model was proved to be more effective than traditional one in the enrichment of professional knowledge and cultivation of comprehensive ability.^2^

The department of periodontology of our school has been built online learning platform for decades. During the current COVID-19 pandemic, many medical schools had to move the offline classes into online platform, and minimize face-to-face practices.^12,13^ In our school, some offline lessons were also postponed or moved online because of epidemic prevention principle, and there is an urgent need to change the traditional learning methods. After thoughtful consideration and discussion, we decided to put this blended learning mode into practice, and to evaluate whether the blended learning mode based on SPOC can achieve a satisfactory effect and get promotion in the future.

## MATERIALS AND METHODS

### Study participants

The students are fourth-year undergraduate students majoring in stomatology of our school (n=65), including 10 overseas students, 5 students under joint-education project from other schools. Before this blended learning experiment, they attended the same prerequisites. The textbook and syllabus they used are the same as those of the previous students. All of the students are volunteered to attend this blended learning project, which is proved and hosted by College of Stomatology, Shanghai Jiao Tong University.

### Optimized the teaching calendar

The teaching calendar was rescheduled, and divided into online and offline part with the same teaching hours as those of the previous teaching calendar. The online classes account for about 20% teaching hours in the form of small private online courses (SPOC), including both theoretical and practice lectures. The content of online courses is chosen carefully and suitable for students to learn by themselves, such as the theoretical knowledge of periodontal anatomy, pathogenic microorganism. Also, practical operations demonstrated online could be watched repeatedly for a better understanding.

### Establishing the online learning platform

The online learning platform was built on the Xfaike online course platform, which is proved by College of Stomatology, Shanghai Jiaotong University. (The online course website is https://shsmu.xfaike.com/classes.). The multimedia materials, including video lectures for theoretical and practice courses, supplementary references, syllabus and so on are available on the Xfaike online course platform for all the students. The relevant multimedia materials are uploaded and open to students 1 week before the class, and the classroom teaching is also recorded and uploaded onto Xfaike online course platform after the class.

### Integration of the online and offline content

The online and offline content of blended learning is tightly integrated by properly designed preview and review parts.

Online pre-learning part is set for some courses, which could help students to preview before the offline class, and guide them to think about the questions arising from online learning. Besides, a quiz is followed after the video lecture to evaluate the result of learning of every student, which is an important feedback for teachers to adjust learning content offline. After the face-to-face class, students need to finish comprehensive questions or take part in discussion after class online for a further understanding of what they learnt previously.

### Evaluation of the feedback

We created our own questionnaire containing 13 questions, and all of the students are volunteered to answer the questions, and the survey was completely anonymous. These questions were divided into 3 parts. The first part was about the practicability of instruction method with 4 questions. The second part was focused on the instruction content with 4 questions, and both theoretic and practical courses were involved. The third part dealt with the efficiency and effectiveness of blended learning mode, containing 5 questions. The questionnaire was designed with a 5-point scale, ranging from 1 point (Strongly disagree) to 5 point (Strongly agree).

### Statistical analysis

The statistical analyses of the data were performed with SAS 9.3 (SAS Institute, Cary, NC, USA). Analysis of variance (ANOVA) and was used to evaluate the data in the different groups. The differences were convinced statistically significant at *p* < 0.05.

### Ethical consideration

The Shanghai Jiao Tong University College of Stomatology institutional review board proved the study and waived the requirement for written informed consent.

## RESULTS

The teaching calendar of periodontology was arranged according the learning content and difficulty of self-learning (Table 1). The theoretical classes on SOPC platform include the introduction, periodontal microbiology and immunology, the relationship between periodontal health and prosthodontic treatment, and so on. For the practical class, the lesson about recognition of the supragingival, subgingival instruments was set online, so students can master basic operation skills via videos. Videos and courseware were open to students a week before the class, and teachers could stay up-to-date with students’ learning progress though management system on website.

**Table 1.** The teaching calendar of periodontology under blended learning mode.

| **Course content** | **Theoretical/Practical** | **Online/Offline** |
| --- | --- | --- |
| **Introduction; Anatomy, structure, and function of the periodontium** | Theoretical | Online |
| **Classification and epidemiology of periodontal diseases** | Theoretical | Offline |
| **Periodontal microbiology and immunology** | Theoretical | Online |
| **Local and host contributing factors of Periodontal diseases** | Theoretical | Online |
| **Clinical features, pathobiology and imaging manifestation of periodontal diseases** | Theoretical | Offline |
| **Gingival diseases** | Theoretical | Offline |
| **Periodontitis** | Theoretical | Offline |
| **Periodontitis as a manifestation of systemic disease** | Theoretical | Online |
| **Other lesions associated with periodontitis** | Theoretical | Offline |
| **Periodontal medicine** | Theoretical | Offline |
| **Assessment of risk factors, prognosis and treatment plan of periodontal diseases** | Theoretical | Offline |
| **The treatment of periodontal diseases** | Theoretical | Offline |
| **Maintenance therapy and prevention of periodontal diseases** | Theoretical | Offline |
| **The relationship between periodontal health and prosthodontic and orthodontic treatment** | Theoretical | Online |
| **Peri-implant tissue and diseases** | Theoretical | Online |
| **Recognition of the supragingival and subgingival instruments and observation of operation skills** | Practical | Online |
| **Periodontal examination, plaque staining, and medical history writing** | Practical | Offline |
| **The operation of hand supragingival scaling, ultrasonic scaling, subgingival scaling and root planning** | Practical | Offline |
| **Gingivectomy, periodontal flap surgery** | Practical | Offline |

Our SPOCs are available on the Xfaike online course platform, which contain abundant learning resources and easily-understood web interface. On the homepage of online classroom, students could easily find the section of courseware, pre-class exercises, discussion board, homework, classroom feedback (Fig 1). The relevant multimedia materials and video recording of offline classes were listed on website according to the order of teaching calendar (Fig 2). The speaking speed of courseware could be adjusted and all of the videos were automatically subtitled, which can help students better understand the content, especially for foreign students.

**Fig 1.**
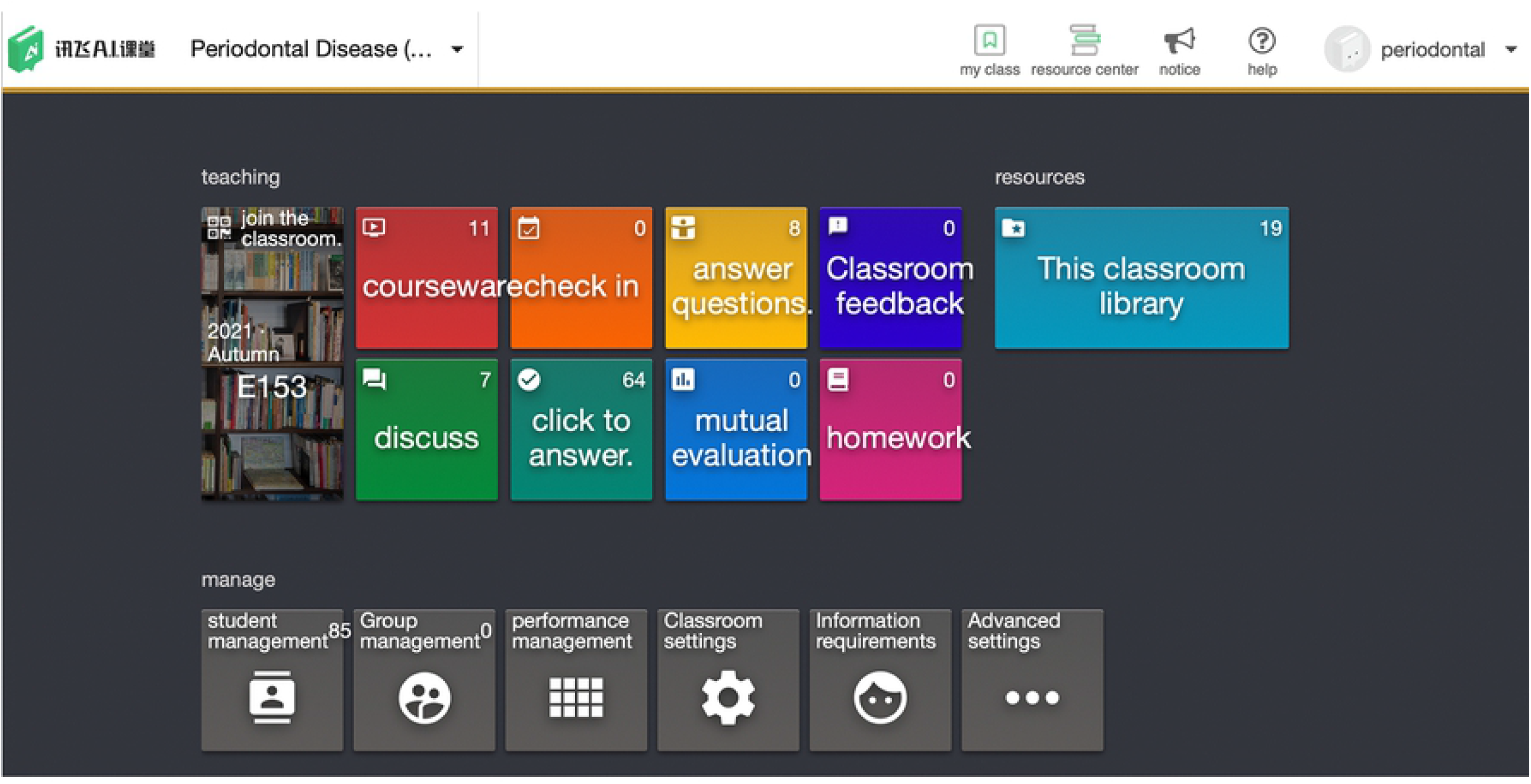
The web page of online courses is built on Xfaike online course platform, which contains videos, courseware homework, etc. Link may be accessed here: https://shsmu.xfaike.com/classes/4153

**Fig 2.**
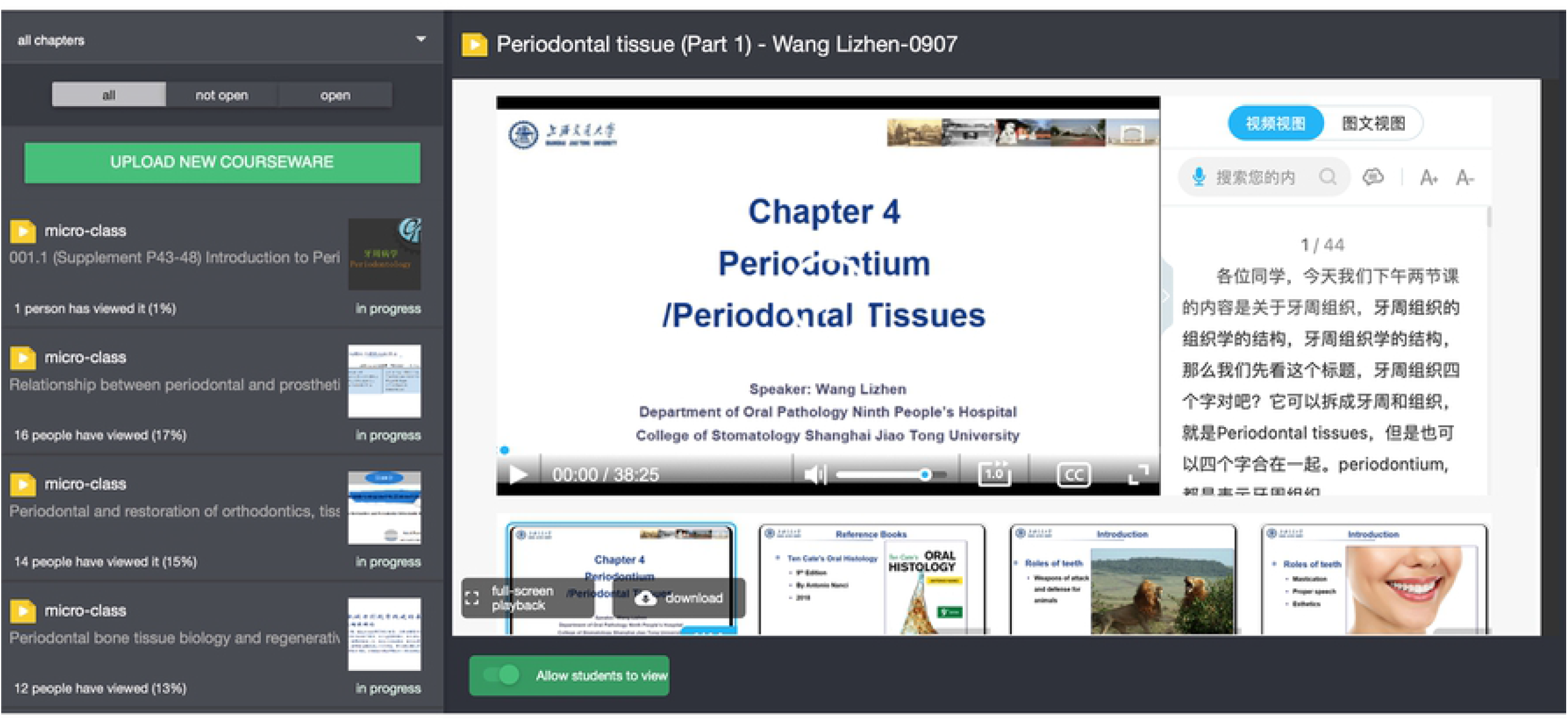
The courseware of online courses. Link may be accessed here: https://shsmu.xfaike.com/classes/2510/coursewares.

All of the students (n=65) answered the questionnaire including 30 males (46.15%) and 35 females (53.85%) (Fig 3). The feedback of blended learning mode were evaluated though three aspects based on the questionnaire we received. According to the feedback of questions in Table 2, the blended learning mode is rather popular and highly accepted among students (Fig 4). About 80% students strongly agreed that they can accept the online and offline blended learning mode and none of them raised objection (4.80/5). 55 students (84.62%) could preview before the offline classes, and think about the questions arising from online learning, and the rest of students (15.38%) failed to preview on time (4.17/5). All of the students believed that they can exchange their questions with teachers more pertinently in offline class after online learning (4.43/5), and online learning videos can be viewed smoothly and repeatedly at any time (4.77/5).

**Fig 3.**
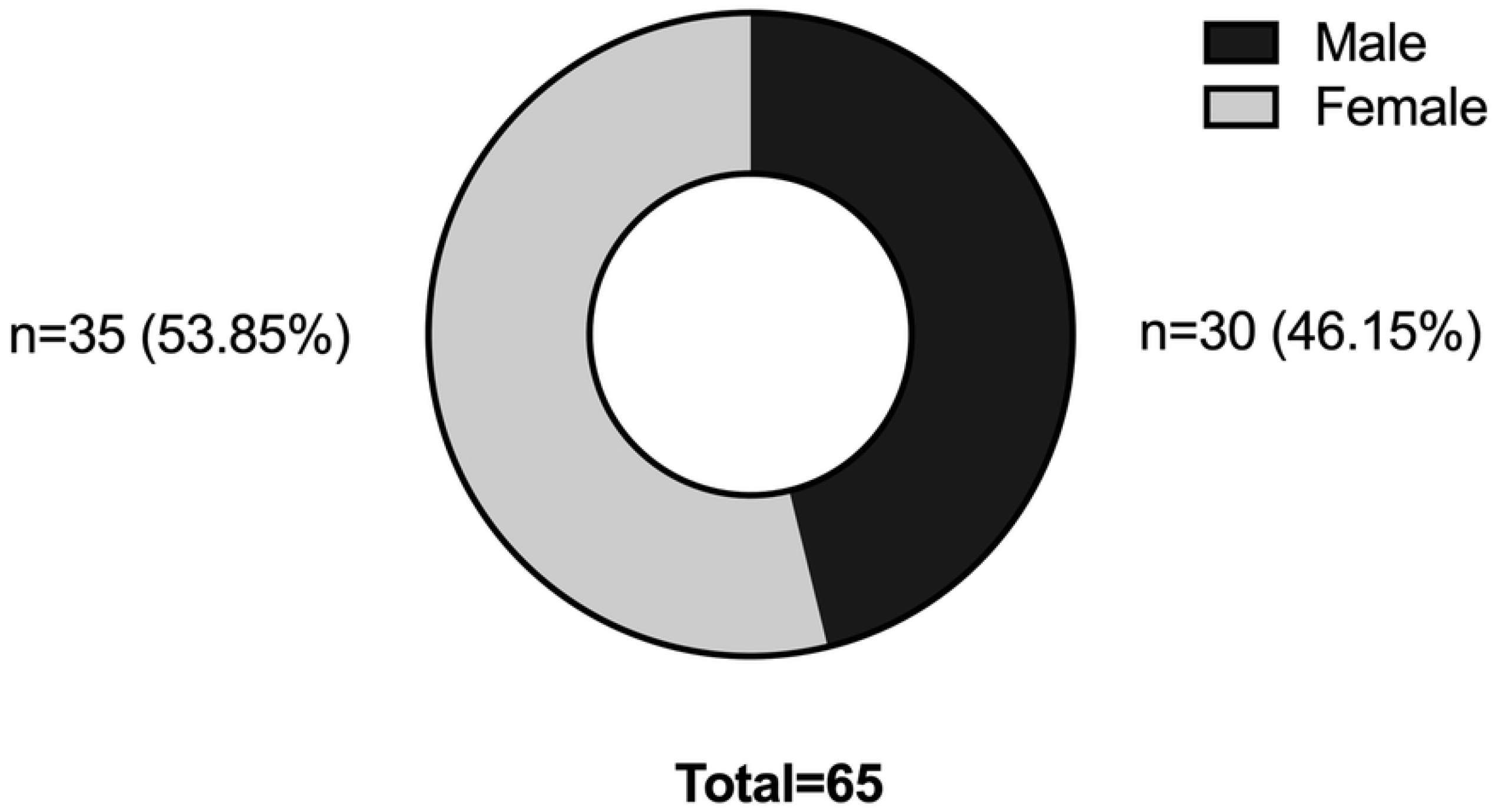
The gender distribution of students who participated in the blended learning.

**Table 2.** Questions about the instruction methods.

| **Questions** | **Mean (SD)\*** |
| --- | --- |
| Q1: Some professional courses were carried out online. I can adapt to this online and offline blended learning mode. | 4.80 (0.40) |
| Q2: Online pre-learning part is set for some courses, I can preview before the offline class, and I will think about the questions arising from online learning. | 4.17 (0.67) |
| Q3: After online learning, I could exchange my questions with teachers more pertinently in offline class. | 4.43 (0.5) |
| Q4: Online learning videos can be viewed smoothly and repeatedly at any time. | 4.77 (0.42) |

5-point Likert scale: 5-Strongly agree, 4-Agree, 3-neither agree nor disagree, 2: disagree, and 1: strongly disagree.
\*SD: standard deviation.

**Fig 4.**
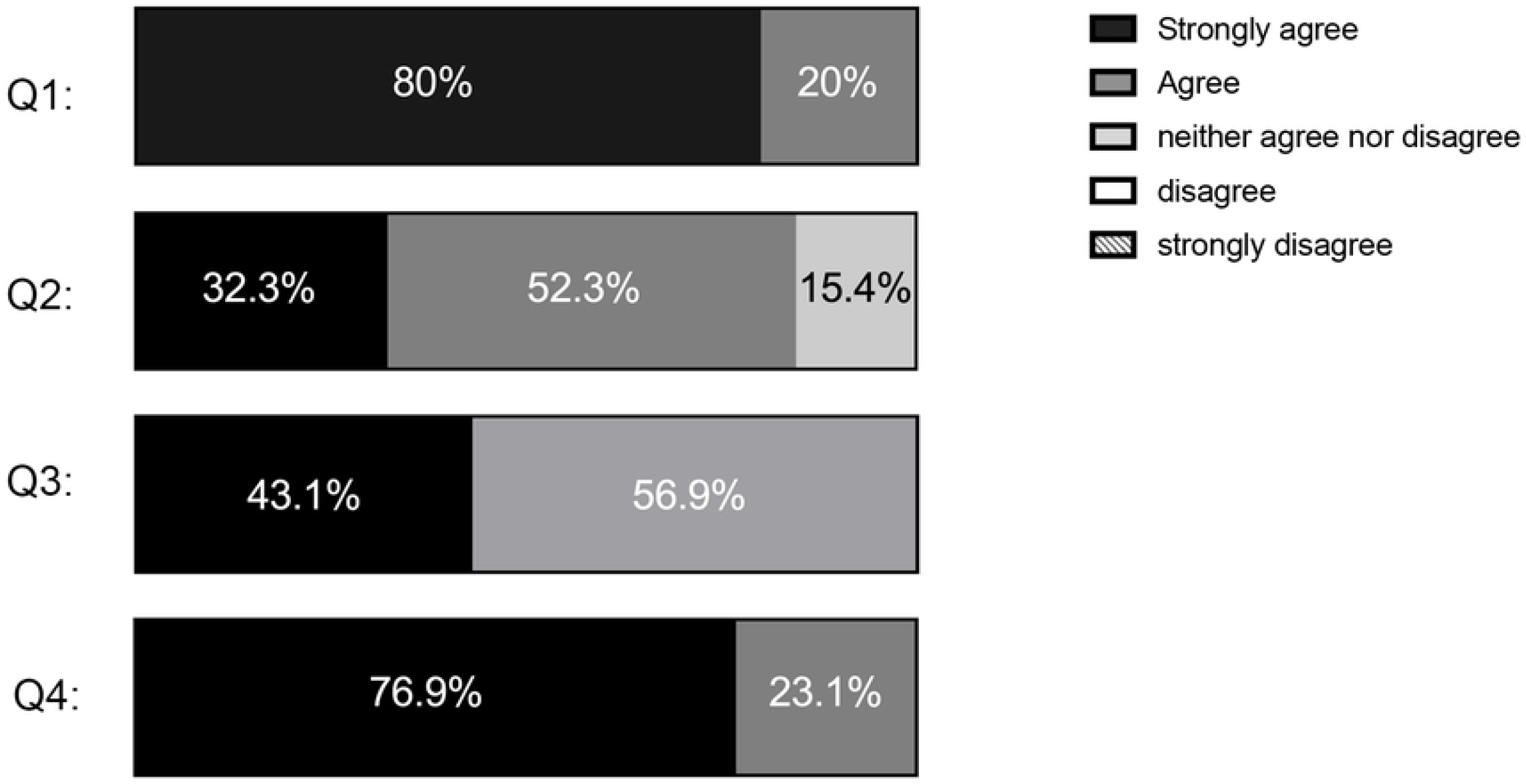
The feedback of blended learning mode on instruction methods.

As shown in Fig 5 and Table 3, 52.3% students strongly agreed that online courses provide abundant learning materials, which can meet their theoretical and practical learning needs, and the others (47.7%) are also agreed with it (4.52/5). What’s more, students are satisfied with the implement of online courses, as the speaking speed and subtitles could be adjusted according to personal habits (4.68/5). Compared with the traditional operating demonstration in the offline classroom, the operation display online is more popular (4.62/5), as the operating steps can be observed more closely, students can benefit more from videos. However, there are 2 students who preferred the face-to-face demonstration. More than 80% students watched online teaching videos for 1-3 times, and 15.4% of students watched the videos for 4-5 times (3.09/5).

**Fig 5.**
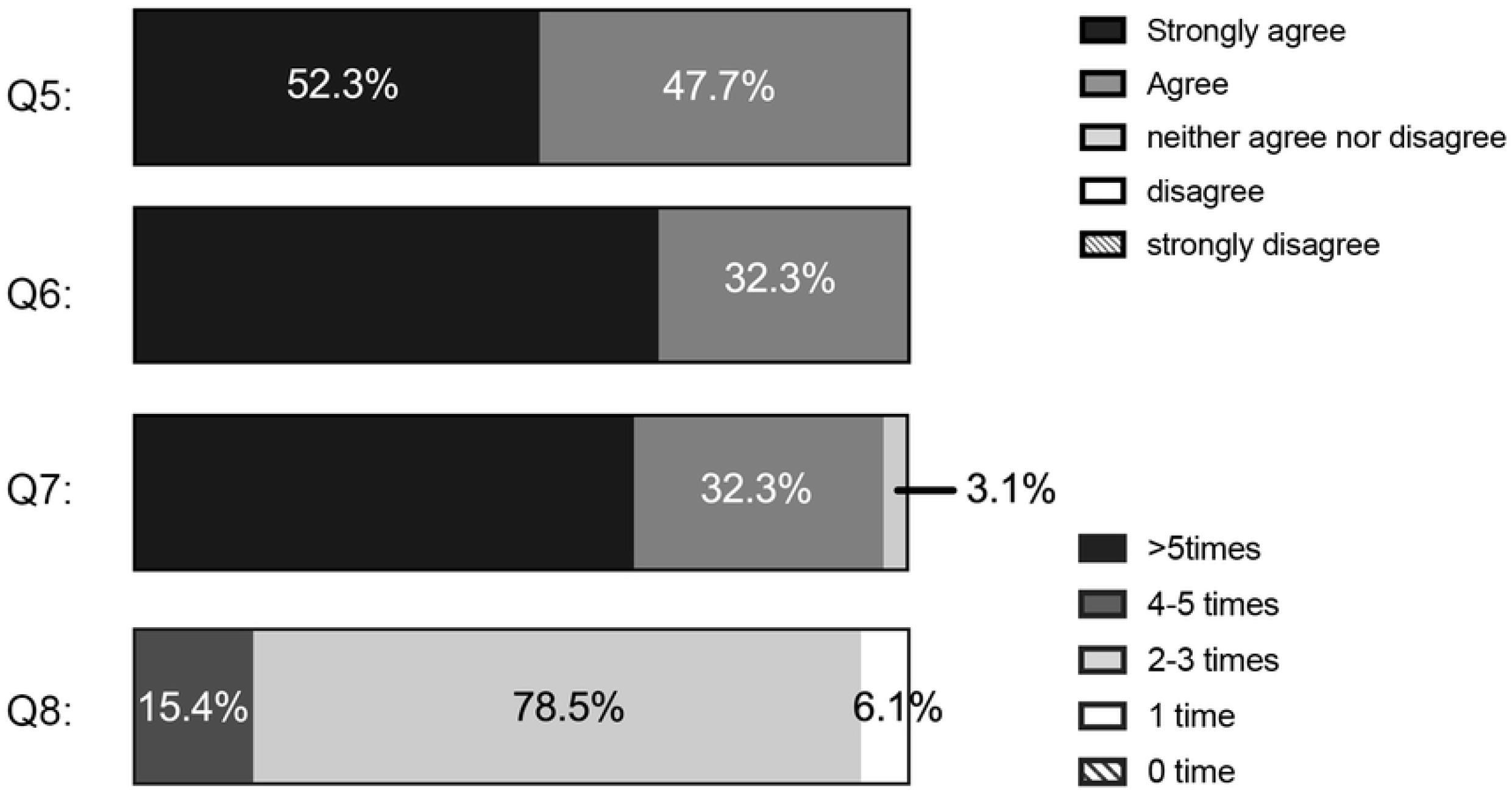
The feedback of blended learning mode on instruction content.

**Table 3.**
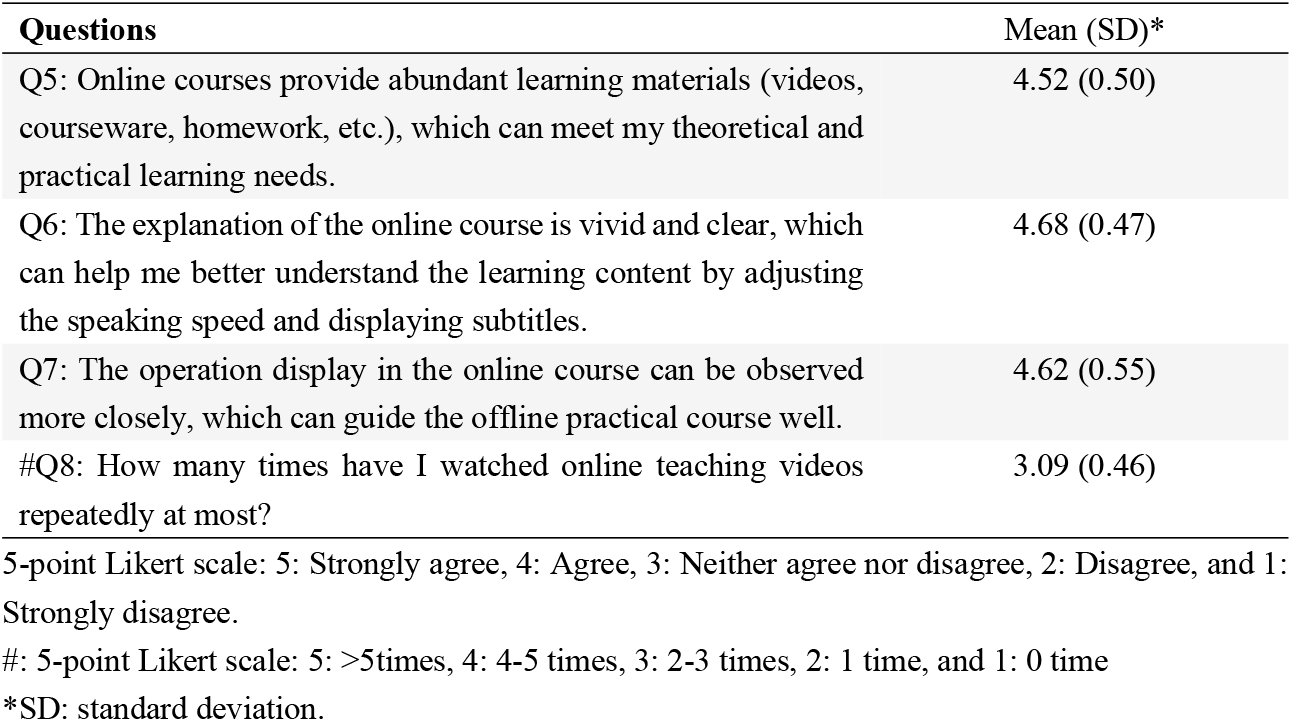
Questions about the instruction content.

When it comes to the efficiency and effectiveness of blended learning mode (Table 4 and Fig 6), more than 95% students convinced that blended learning mode can improve their self-learning ability (4.63/5), and about 85% students were inspired and guided by the supplementary reading materials, such as the latest scientific research progress in the specialization (4.32/5). However, there are different opinions on the difficulty of online exercises. About 80% students agreed that difficulty of exercises and topics for discussion was moderate, and completion of exercises and discussion could help them better preview and review the contents of subsequent classes (4.23/5) and have deeper understanding of the knowledge (4.38/5). Meanwhile, a small number of students complained that some of the exercises and topics were too hard for them to answer. Despite of the deficiencies that need to be improved, all of the students can adapt to the blended learning mode with higher proportion of online courses (4.57/5).

**Table 4.** Questions about the efficiency and effectiveness of blended learning mode.

| Questions | Mean (SD)* |
| --- | --- |
| Q9: Blended learning mode can improve my self-learning ability. | 4.63 (0.55) |
| Q10: The addition of scientific research progress in blended learning courses could inspire and guide me, and mobilize my learning enthusiasm. | 4.32 (0.73) |
| Q11: The difficulty of online exercises is moderate, which can help me better preview and review the contents of subsequent classes. | 4.23 (0.88) |
| Q12: After class discussion on the online platform can help me have a deeper understanding of the content of offline classes. | 4.38 (0.68) |
| Q13: If the proportion of online courses is raised according to teaching content, I can adapt to this blended learning mode. | 4.57 (0.50) |

5-point Likert scale: 5-Strongly agree, 4-Agree, 3-neither agree nor disagree, 2: disagree, and 1: strongly disagree.
\*SD: standard deviation.

**Fig 6.**
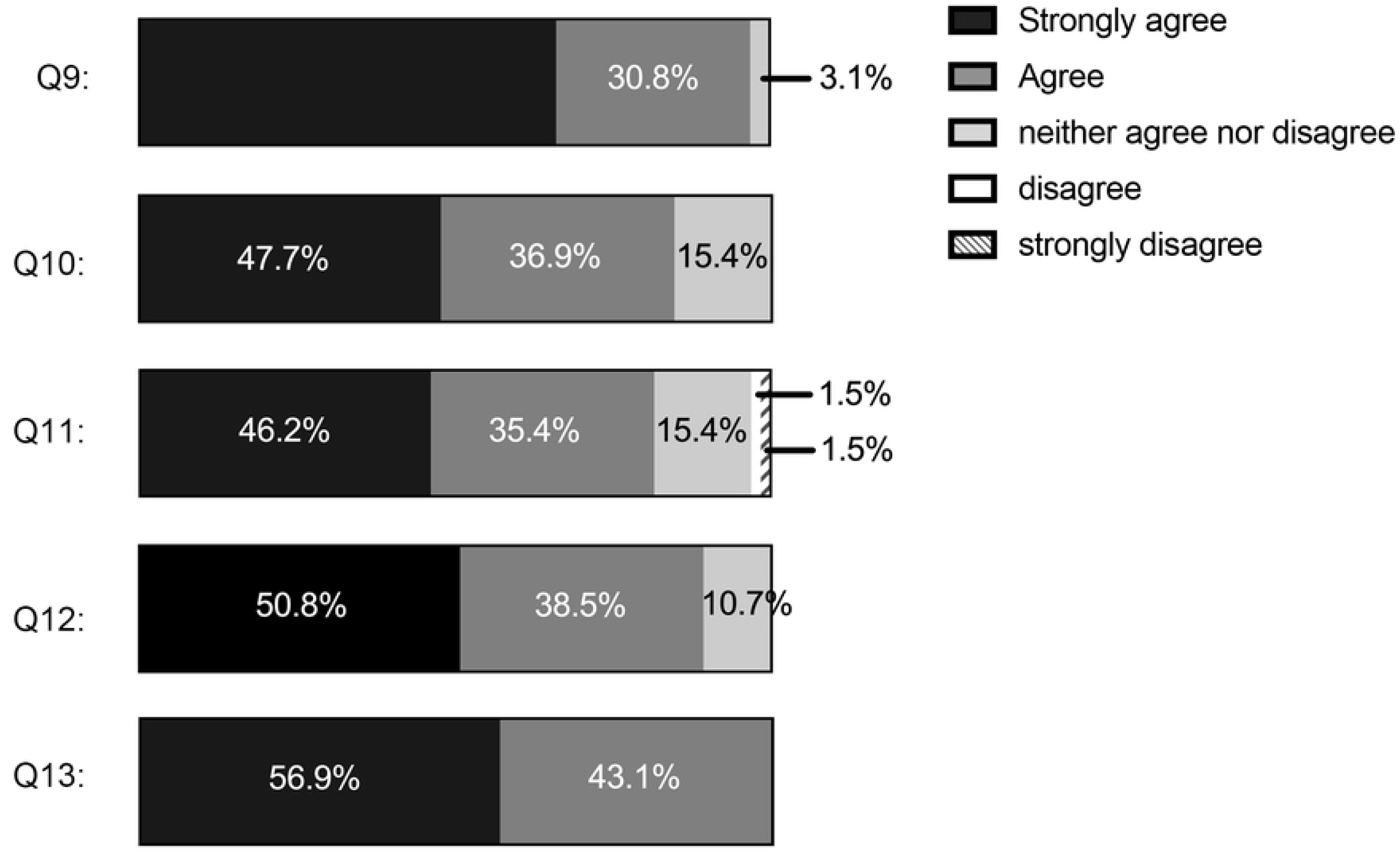
The feedback on the efficiency and effectiveness of blended learning mode.

## DISCUSSION

This study is aimed to investigate the learning effect of blended learning mode, consisted of SPOC and offline classes, for the further optimization and reform in the medical education. Considering the COVID-19 pandemic, some of the medical school postponed or suspended the classes. In order to maintain the continuity and quality of medical education, this blended learning mode was carried out in fourth grade students of Stomatology who began to learn major courses.

In this blended learning platform, we prepared abundant learning content online, including video, courseware, and references. The teaching procedures were properly designed, and preview and review parts were assigned to integrate the online and offline parts tightly. After the whole project, we collected and organized students’ opinions toward the instruction method, content and achievements during their learning process. According to the questionnaire, blended learning mode has been widely accepted by students and they hold positive attitude toward a higher proportion of online course in the future.

Although all of the students are satisfied with the instruction content, we still received comments and suggestions from the teaching staff. For example, medical ethics education should be emphasized, and the syllabus will be modified subsequently with more attention on the humanistic concern. The courseware will be re-recorded with uniform format to ensure the quality, as some of the videos are the recording of offline class previously. Besides, the management of online courses platform is vital to the blended learning mode. The communication and interaction between students and teachers needs to be enhanced online, a fixed time would be set aside every week to answer students’ questions.

The online exercises and after class discussion also need modification. The questions and topic for discussion could be more inspiring and interesting, and students are allowed to work in small teams to accomplish the tasks in the future. The classroom teaching could also influence the learning effect of SPOC. As the classroom is still the main part of the program, teachers in face-to-face class could provide more support to the students who have difficulties in the online courses.

In conclusion, we applied the blended learning mode in the theoretical and practical education of periodontology. The blended learning platform is practicability, and the learning effect is promising, but there are still some shortcomings in both SPOC and offline class that need to be improved.

## CONCLUSION

Nowadays, some practical on blended learning mode in medical education have been carried out, however, there is few study about the feasibility and learning effect on dental education. Our study performed this blended learning mode on the stomatological students in our school under COVID-19 pandemic, and received promising feedback. Hence, it is concluded that the blended learning mode is a practical way to improve the learning effectiveness and efficiency for students, which is worth further promotion.

## Data Availability

Data cannot be shared publicly because of ethical considerations. Data are available from the Shanghai Jiao Tong University College of Stomatology Institutional Data Access / Ethics Committee (contact via Tel:86 021-23271699) for researchers who meet the criteria for access to confidential data.

